# Racial Bias Among Doctor of Physical Therapy Students and its Impact on Managing Patients in Pain

**DOI:** 10.1101/2025.04.13.25325754

**Authors:** Marlon L. Wong, Neva Kirk-Sanchez, Elizabeth A. R. Losin, Gregory W. Hartley

## Abstract

**Purpose/Hypothesis:** The purpose of this study was to determine 1) the prevalence of false beliefs about biological differences between Blacks and Whites among Doctor of Physical Therapy (DPT) students, and 2) if DPT students evaluate and approach the management of pain differently based on the race of a hypothetical patient.

**Methods:** DPT program directors across the United States were emailed and asked to distribute the survey to students currently enrolled in their respective DPT programs. The survey consisted of 15 questions on beliefs of biological differences between races, as well as questions on the estimated pain intensity and treatment parameters for gender-matched patient cases (one with an ankle sprain and another with low back pain) with randomized race.

**Results:** Of 457 respondents, 72% were female and 80% were White non-Hispanic. Respondents came from 47 states, and 27% of respondents reported growing up in rural, 63% in suburban, and 10% in urban communities. 71% of respondents held at least one false belief about biological differences between Black and White people. Students appraised the hypothetical Black patients with slightly higher pain and recommended more treatment for the Black patients (p<0.05). There were no differences in pain appraisal between students who did and did not hold false beliefs.

**Conclusions:** The majority of students had misconceptions regarding biological differences between Black and White races. However, these misconceptions did not result in meaningful differences between how DPT students appraised or proposed to manage hypothetical Black and White patients.

**Clinical relevance:** Less than 5% of physical therapists in the United States are Black, and racial bias among physical therapists has not been widely studied. By improving understanding of bias and false beliefs in DPT students, this study provides valuable information on the readiness and barriers for our future workforce to provide equitable care.

## INTRODUCTION AND BACKGROUND

Musculoskeletal pain conditions are a major challenge for the United States (US) health care system, with a 2019 National Health Interview Survey revealing that 58.9% of adults over the age of 18 had experienced musculoskeletal pain in the past 3 months (1). Further, musculoskeletal pain conditions are associated with the highest economic costs among common conditions requiring rehabilitation in the US (2), costing the nation an estimated $360.0-$405.4 billion per year (3). Clearly, training health care providers to meet these pain management needs is critical. However, racial health inequities are well documented in the US, and they are a barrier to meeting societal needs for improved pain management (4). This is exemplified by the finds of Khosla, et al. (5). that “American clinicians displayed less optimistic expectations for the medical treatment and health of a Black male patient, relative to a White male patient, and that this racial bias was related to their view of the Black patient as being less personally responsible for his health relative to the White patient.” In fact, studies have repeatedly shown that pain is underestimated and undertreated in Black patients compared to White patients (6–8).

Given the importance of training future health care professionals to meet the rising demand for pain management, and the fact that racial bias is a known barrier to achieving this goal, it is also important to assess and intervene on provider bias within their respective training programs. In one of the few studies of its kind, Hoffman et al. (9) surveyed 222 White medical students and residents and found racial bias in pain assessment and treatment recommendations and false beliefs about biological differences between Blacks and Whites. Half of their participants endorsed false beliefs about biological racial differences, and participants who endorsed these beliefs rated the Black (vs. White) patient’s pain as lower and made less accurate treatment recommendations (9).

Lane, et al. (10) studied the differences in pain experience among patients in different racial and ethnic groups. This work provided further support of differences in pain experiences among Black patients and non-Hispanic White patients with chronic spinal pain receiving physical therapy. However, to our knowledge, racial bias in pain assessment and treatment has not been studied in Doctor of Physical Therapy (DPT) students as the providers of care. This is an important gap to address as physical therapy has been advocated for musculoskeletal pain as a non-pharmacologic approach to decrease opioid use (11), and early access to physical therapy has been shown to decrease health care costs and improve patient outcomes (12).

Thus, we aimed to use the Hoffman et al.(9) study as a framework to investigate the presence and impact of false beliefs and racial bias in DPT students. The purpose of this study was to determine if 1) DPT students evaluate and approach the management of pain differently based on the race of the hypothetical patient, 2) the prevalence of false beliefs about biological differences between Blacks and Whites among DPT students, and 3) if there are differences between those DPT students who hold false beliefs and those who do not in how they evaluated and manage of pain in hypothetical Black and White patients.

## MATERIALS AND METHODS

### Participants/Recruitment

Students currently enrolled in DPT programs were recruited in two phases from February 2021 until August 2023. In the first phase, emails were sent to DPT program directors across the state of Florida asking them to distribute the survey to their student body. In the second phase, program directors across the nation were emailed, flyers were distributed at two national conferences (the American Physical Therapy Association Combined Sections Meeting and the Education Leadership Conference), and information on the study was distributed via the American Council of Academic Physical Therapy website. All participants consented electronically prior to initiating the survey, and all data were analyzed anonymously. To provide electronic consent, participants read the online survey consent script and then were presented with the prompt *“By selecting [“I consent to participate”] below, this means you consent to participate in this research project.”* The University of Miami Institutional Review Board provided approval for this study and the consent process (# 20201416).

### Survey

The survey was modelled after a study conducted by Hoffman et al.(9) on the racial bias of medical students. However, for this study, the survey was modified to be more meaningful and relevant to the physical therapy profession. For example, the cases used by Hoffman et al.(9) involved a patient with a fractured bone and a patient with a kidney stone, and these cases required participants to recommend medications for pain management. Conversely, the patient cases used in our study involved patients with low back pain and an ankle sprain and required participants to recommend treatment intensity (duration and frequency).

After consenting, participants were asked to confirm their status as a current student in a physical therapist education program and to provide their age and gender. The survey program would then route the participant to gender-matched patient cases but with randomized race (i.e., Black or White). The order of patient race was counterbalanced across participants. Each participant was exposed to two gender-matched patient cases: the first case was of a person with “acute low back pain with radiating pain associated with disc herniation”, and the second case involved a person with “acute lateral ankle sprain (grade IIIB).” These conditions were chosen due to the high frequency in which physical therapists typically encounter these conditions. Low back pain is among the most prevalent health conditions in the United States, with 70% of people experiencing spinal pain symptoms at least once in their lifetime (13, 14). Ankle injuries are also very common, with approximately 30,000 ankle injuries occur every day in the United States, and an incidence rate estimated at 2.15 per 1000 persons (15).

Each case contained a brief description of the primary complaint and mechanism of injury, physical examination findings and imaging results, and the medical diagnoses listed above. Identical to the study by Hoffman et al.(9), we manipulated the patient’s race in 2 ways: stereotypical Black or White names were used at the top of the case description (i.e., (Black names: Taneisha, Kiesha, Darnell, Jermaine; White names: Hannah, Katelyn, Brett, Connor), and the patient’s race was provided within the case description (i.e., African American or Caucasian male/female). For all patient cases, participants were asked to estimate the patients’ pain intensity, suggest a frequency and duration of physical therapy treatment.

In the second part of the survey, participants provided information on their race, ethnicity, nationality, State of residence, type of community (i.e., rural, suburban, or urban), and current semester in physical therapy school (e.g., third semester).

The final part of the survey required participants to rate 15 statements about biological differences between Black and White people that are true or untrue on a six-point scale (1 = definitely untrue, 2 = probably untrue, 3 = possibly untrue, 4 = possibly true, 5 = probably true, 6 = definitely true). Of the 15 items, 4 were true and 11 items were false statements. Of the false statements, 8 were in the direction of Black strength and White frailty and 3 were in the direction of White strength and Black frailty.

### Analysis

Surveys were considered complete if respondents answered questions related to pain intensity and treatment parameters, and incomplete surveys were removed from the sample. Data with outliers related to treatment duration and intensity were deleted from the sample. This included responses about treatment intensity that was greater than 50 sessions. Descriptive statistics were used to characterize the sample according to year of study, gender, and self-reported race and ethnicity.

Respondents were categorized as high bias if they held at least 2 false beliefs about biological racial differences, and low bias if they held 0 or 1 false belief. T-tests were used to compare pain appraisal and recommended treatment parameters for the back pain and ankle sprain patients between high bias and low bias respondents. T-tests were also used to compare pain appraisal and treatment parameters for Black vs White patients, and 2X2 analysis of variance (ANOVA) was used to compare the main effects for bias category, patient race, and the interaction between the two for both back pain and ankle sprain patients. Finally, multivariate regression analyses were used to test models to predict treatment intensity based on pain appraisal, bias, and patient race.

## RESULTS

Seven hundred twelve students initiated the survey, with 472 students completing it and included in the analyses: 37.7% were first year students, 37.3% were second year students, and 25.0% were third year students. **Figure 1** shows that there were lower proportions of Black/African American and Asian respondents, a similar proportion of Hispanic/Latino respondents, and a higher proportion of White/non-Hispanic respondents as compared to the proportion of physical therapist graduates from those racial/ethnic groups. Respondents represented all regions of the US (**Figure 2**) and the percentage coming from rural vs suburban vs urban environments can be found in **Table 1**. Most respondents both lived in and were raised in suburban environments. When responding to the questions about beliefs about Black versus White people, 71% held at least one inaccurate belief, and 24% held four or more inaccurate beliefs. The percentage of people holding a false belief about each question is found in **Figure 3**, where “yes” indicates a false belief. There were no differences in bias scores or bias categories between first, second-, and third-year students.

**Figure 1.**
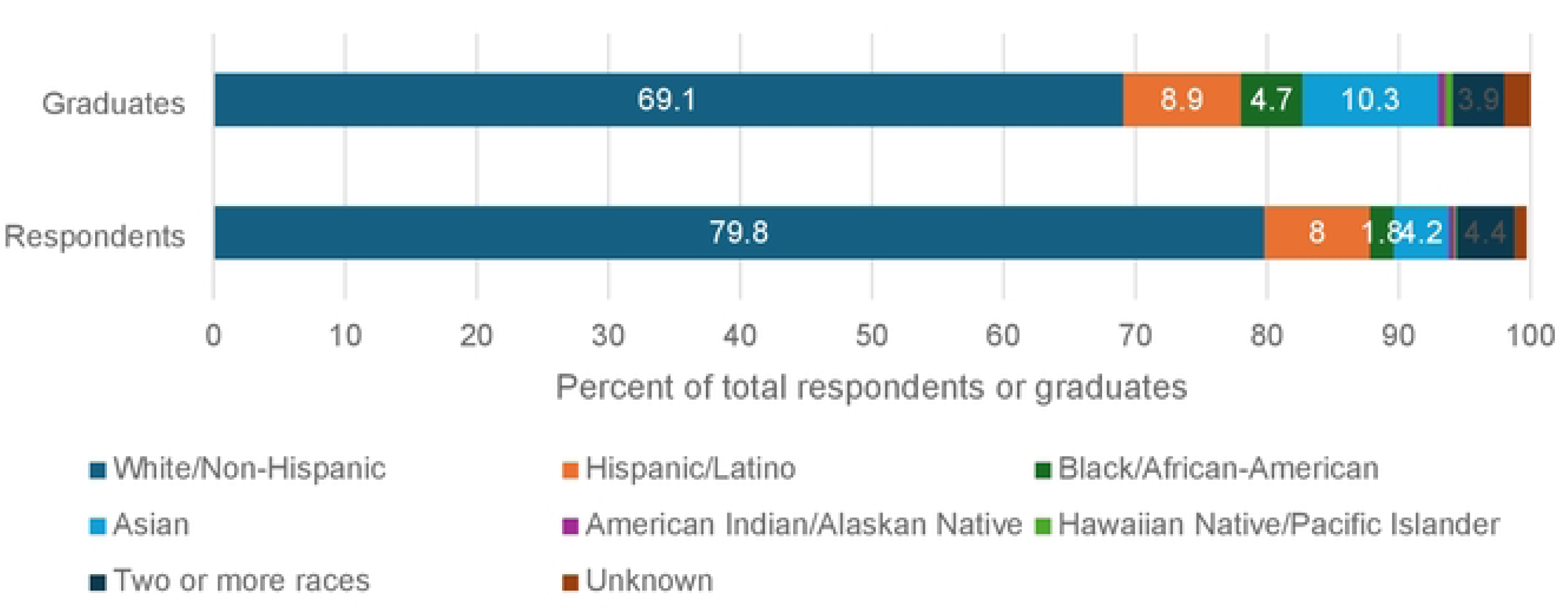
Comparison between race/ethnicity of respondents vs United States graduates

**Figure 2.**
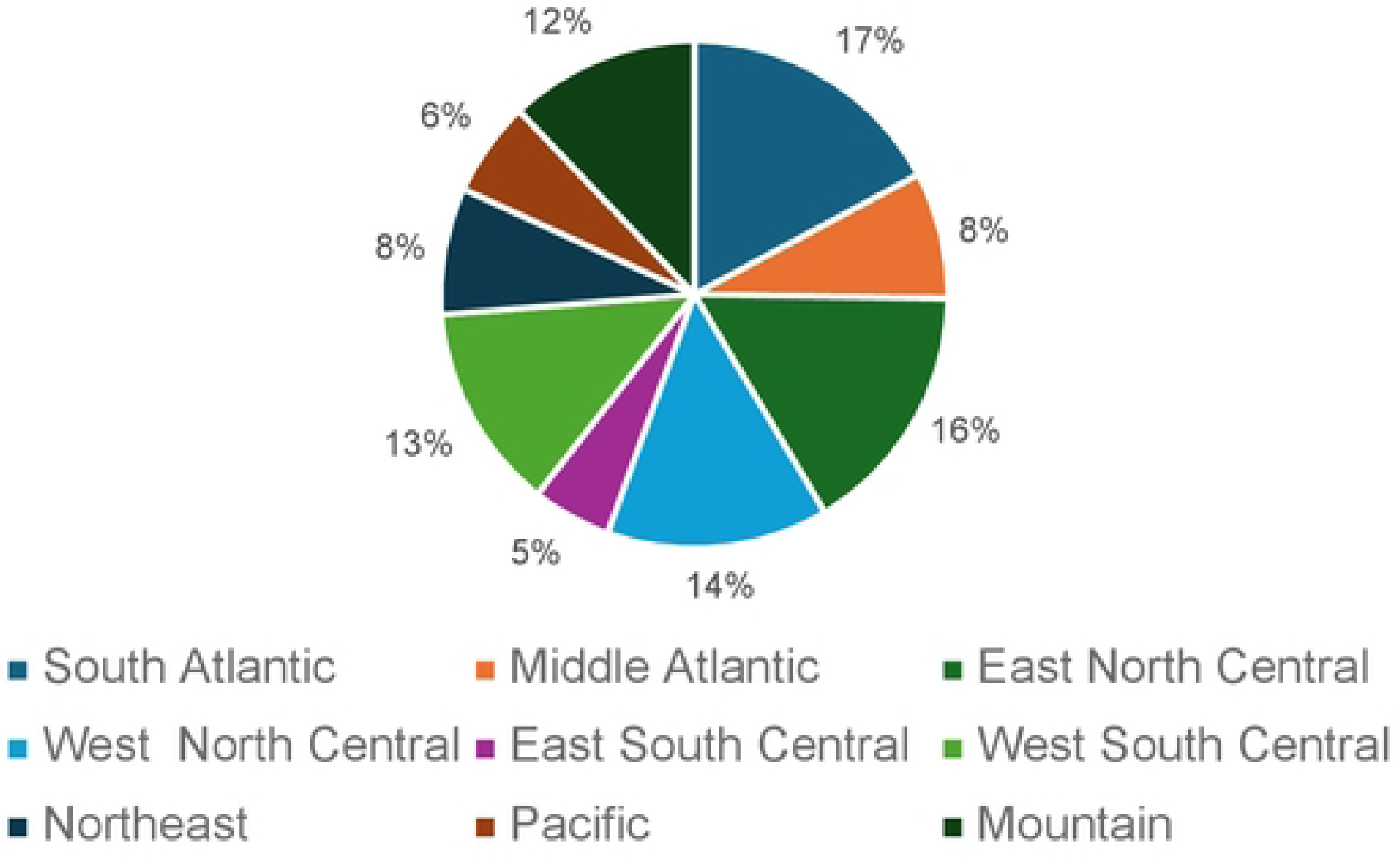
Percentage of respondents by region

**Figure 3.**
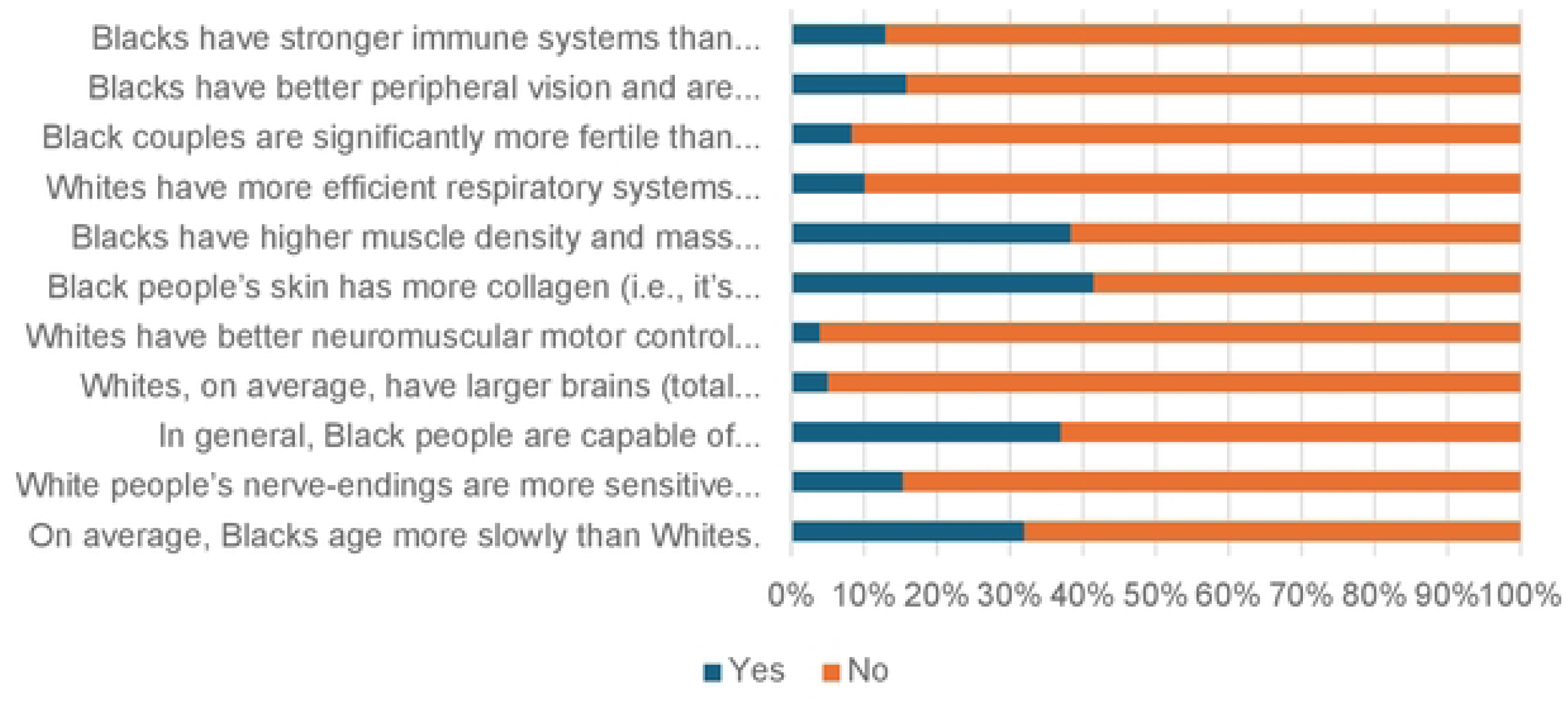
Percent of respondents answering questions incorrectly reflecting false beliefs about Black people

**Table 1:**
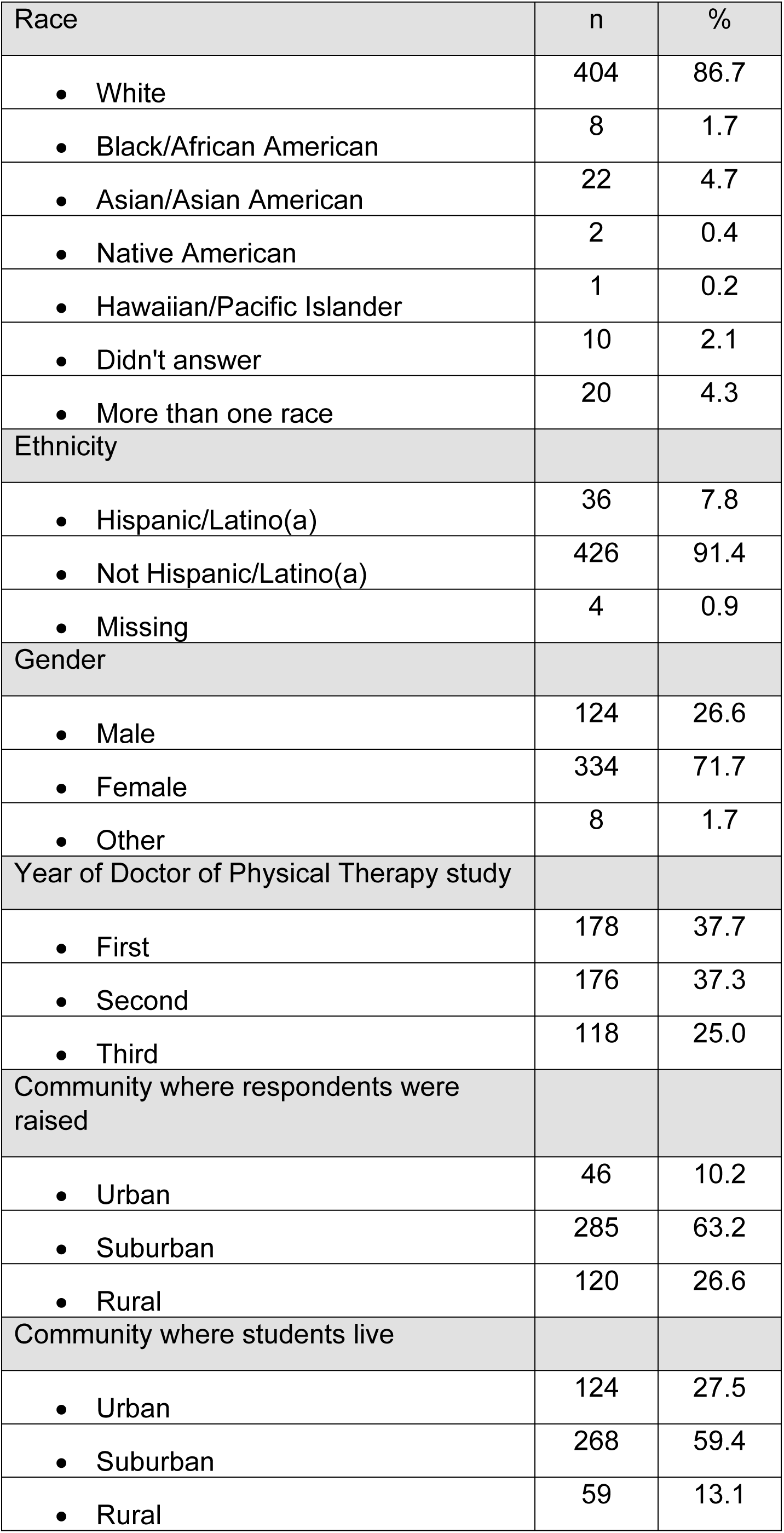
Respondent demographics.

| Race | n | % |
| --- | --- | --- |
| • White | 404 | 86.7 |
| • Black/African American | 8 | 1.7 |
| • Asian/Asian American | 22 | 4.7 |
| • Native American | 2 | 0.4 |
| • Hawaiian/Pacific Islander | 1 | 0.2 |
| • Didn't answer | 10 | 2.1 |
| • More than one race | 20 | 4.3 |
| Ethnicity |  |  |
| • Hispanic/Latino(a) | 36 | 7.8 |
| • Not Hispanic/Latino(a) | 426 | 91.4 |
| • Missing | 4 | 0.9 |
| Gender |  |  |
| • Male | 124 | 26.6 |
| • Female | 334 | 71.7 |
| • Other | 8 | 1.7 |
| Year of Doctor of Physical Therapy study |  |  |
| • First | 178 | 37.7 |
| • Second | 176 | 37.3 |
| • Third | 118 | 25.0 |
| Community where respondents were raised |  |  |
| • Urban | 46 | 10.2 |
| • Suburban | 285 | 63.2 |
| • Rural | 120 | 26.6 |
| Community where students live |  |  |
| • Urban | 124 | 27.5 |
| • Suburban | 268 | 59.4 |
| • Rural | 59 | 13.1 |

Students were categorized as minority if they were non-White or Hispanic. When comparing inaccurate beliefs by minority status, there was no significant difference in the proportion of minority vs non-minority respondents who held at least one inaccurate belief (69% of non-minority and 78% of minority respondents, p for chi-square=0.11).

Differences in pain appraisals and recommended treatment intensities for Black vs White patients are found in **Figure 4**. For the ankle sprain condition, students appraised Black patients with more pain (7.3 vs 6.7, p<0.0001), and recommended higher treatment intensity (17.3 vs. 15.4 hours, p=0.02). Similarly, for the back pain condition, students appraised Black patients with more pain (7.8 vs 7.0, p<0.001) and recommended higher treatment intensity (19.8 vs 15.4 hours, p<0.0001). ANOVA comparing responses of high bias (≥2 false beliefs) and low bias students for Black and White patients with ankle sprain found no main effect for student bias and no interaction effect (F=1.74, p= .15). For patients with back pain, there was a main effect for student bias with high bias students appraising pain level lower than low bias students, regardless of race (least square means 7.2 and 7.5 respectively, p=0.01) but no interaction between patient race and student bias.

**Figure 4.**
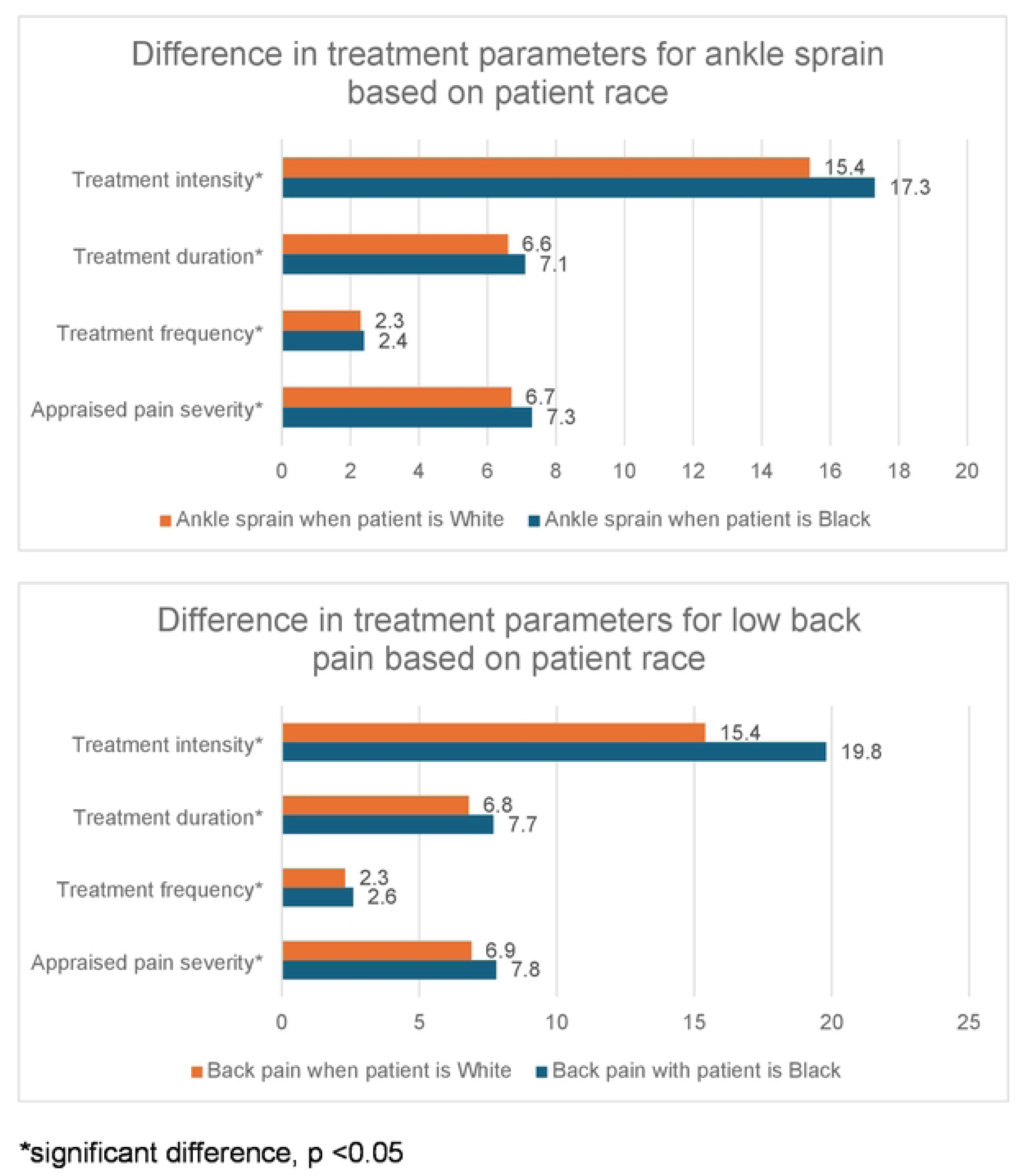
Difference in treatment parameters for ankle sprain and back pain based on patient race

In all cases, appraised pain severity was weakly correlated with recommended treatment intensity, and these correlations were stronger for back pain than ankle sprain patients (**See Table 2**). There were no differences between students with high and low bias for pain appraisal and recommended treatment parameters, except that students with high bias rated White patients with back pain lower than students with no bias. (mean ± standard deviation of 6.2 ± 1.6 for high bias respondents vs 7.1 ± 1.5 for no bias respondents (p=0.005)). The regression model to determine predictors of treatment intensity controlling for bias, race of patient, and appraised pain, was significant for both the ankle sprain (F=18.26, p<0.0001) and low back pain (F=11.08, p<0.0001); however, the R-square values were very low (0.07 and 0.11, respectively), indicating that the models only account for 7% of the variance in treatment intensity for the ankle sprain case (6% by severity and 1% by patient race) and 11% of the treatment intensity in the low back pain case (4% by severity and 7% by patient race).

**Table 2:**
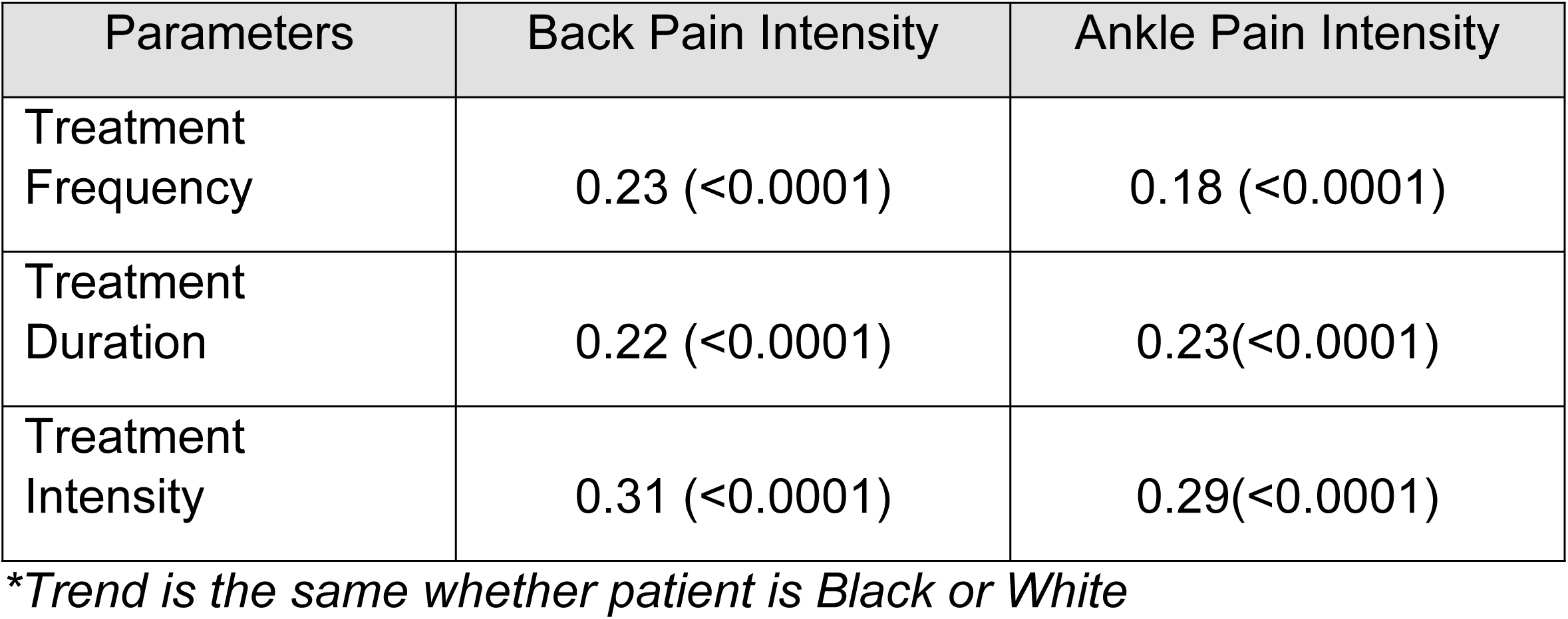
Spearman correlation between appraised pain intensity and treatment parameters [r value (p value)].

## DISCUSSION

In this national survey study of DPT students, there were widespread misconceptions regarding biological differences between Black and White races; however, these misconceptions did not result in meaningful differences between how DPT students appraised or proposed to manage hypothetical Black and White patients with the same condition. Overall, DPT students provided similar estimates of pain intensity and proposed similar treatment plans irrespective of patient race. In fact, the Black patients were rated with slightly higher pain and prescribed more intensive therapy, although the differences were small.

The fact that a high rate of false beliefs about biological racial differences was not associated with meaningful differences in the appraised pain, or the proposed physical therapy management, calls into question the relationship between beliefs and biased behavior in this context. Several philosophers have argued that implicit biases are beliefs (16–18), and that that the dissociations observed using explicit measures likely arise from deliberate judgment processes (19). Others have argued that the instability of implicit biases (i.e., low retest reliability(20) and low correlations between measures of bias(21, 22)), and the fact that biases often do not change in response to changes in evidence, are indicative that biases are not beliefs. While an in-depth discussion on these philosophical underpinnings is beyond the scope of this manuscript, it is important to note that bias and belief are complex constructs, that there is ongoing debate and research on the relationship between the two, and that beliefs can be fragmented and only sometimes spontaneously guide actions, thoughts, and feelings (23).

Surprisingly, students appraised the pain of Black patients higher than White patients in both ankle sprain and back pain cases; congruently, they also prescribed higher treatment intensity for Black patients. This finding conflicts with evidence that racial bias in practitioners often contributes to the undertreatment of pain in Black patients (24–26). However, we used a test to detect differential patterns in *deliberative* reasoning rather than spontaneous rapid behavior (e.g., The Implicit Association Test) to estimate implicit biases. Thus, it is possible that student participants were aware of their own biases and adjusted their responses by overcorrecting for their biases. It is also important to note that, drastic shifts in societal views occurred between the time of Hoffman et al.’s findings (2016) and when this study was initiated in 2021. Importantly, 2020 was a pivotal moment in America’s history, with a widespread push for racial justice after the COVID-19 pandemic disproportionately affected racial minority communities and several high-profile cases of police brutality against Black people incited social unrest. Given the depth and breadth of coverage on racial inequities at this time, it is not surprising that less explicit bias (i.e., relative undertreating pain for the Black patient) was noted in this study compared to Hoffman et al.’s findings from 2016. Further, recent evidence by Lane et al. (10) supports the assertion that there are racial differences in the pain experience among patients receiving physical therapy, with Black patients who have chronic spinal pain reporting higher pain catastrophizing and lower self-efficacy scores at baseline (10). Thus, personal, cultural, environmental, and social differences influence the individual pain experience and racial differences can and do exist. An increased awareness about these differences could impact the perceptions of healthcare providers.

The findings in this study provide unique data as there have only been a few studies on racial bias among physical therapists. Similar to our findings, Cavanaugh and Rauh(27) found no significant difference in treatment frequency for a hypothetical Black woman and a hypothetical White woman with arthritis in a 2021 survey study (27). However, this study was limited to 83 participants (licensed therapists and students) in a single Southern California county (27). Conversely, our findings are based on wide geographic and community settings, with respondents from all major geographic regions in the U.S. and from rural to urban communities.

### Limitations

The survey used in this study was derived from a similar study in medical students; however, the findings in this study should be interpreted with caution given that the survey has not been validated. Additionally, there was underrepresentation of some minoritized groups, which may affect generalizability of our findings. Nevertheless, our sample is consistent with the underrepresentation found within the physical therapy field. In the physical therapy workforce, people who self-identify as Black (4.2%), Hispanic/Latino(5.4%), or two or more races (1.3%), make up approximately 11% of practicing therapists.(28) Similarly, these groups made up approximately 14% of our sample.

### Conclusion

The majority of DPT students had misconceptions regarding biological differences between Black and White races. However, these misconceptions did not result in meaningful disparities between how DPT students appraised or proposed to manage hypothetical Black and White patients. It is important to note that currently less than 5% of physical therapists are Black, and racial bias among physical therapists has not been widely studied. By improving understanding of bias and false beliefs in DPT students, this study provides valuable information on the readiness and barriers for our future workforce to provide equitable care.

## Data Availability

All relevant data are within the manuscript and its Supporting Information files.

